# 2023-24 dengue outbreak in Valle del Cauca, Colombia caused by multiple virus serotypes and lineages

**DOI:** 10.1101/2024.07.16.24310413

**Authors:** Nathan D. Grubaugh, Daniela Torres-Hernández, Mónica A. Murillo-Ortiz, Diana M. Dávalos, Pio Lopez, Isabel C. Hurtado, Mallery I. Breban, Ellie Bourgikos, Verity Hill, Eduardo López-Medina

**Author notes:** Address for correspondence: Nathan D. Grubaugh, Yale School of Public Health, 60 College St, Suite 608, New Haven, CT 06510, USA.

## Abstract

Global dengue cases rapidly rose to record levels in 2023-24. We investigated this trend in Valle del Cauca, Colombia to determine if specific dengue virus serotypes or lineages were responsible for the large outbreak. We detected all four serotypes and multiple lineages, suggesting that other factors, such as climatic conditions, are likely responsible.

## Introduction

Reported cases caused by dengue virus (DENV: *Flaviviridae*; *Orthoflavivirus*), composed of four genetically distinct serotypes (DENV-1-4), are on the rise. In 2023, 4.6 million dengue cases were reported, a record at the time and a 64% increase over 2022 (1). Those numbers were quickly surpassed in 2024 with almost 10 million dengue cases through June. Of those, ∼8.4 were reported from Brazil alone; however, large outbreaks have been reported by many countries, including Colombia. The Valle del Cauca State Health Department in Colombia reported ∼56,000 dengue cases through May, 2024, compared to ∼23,000 for all of 2023 and less than 5,000 in 2022 (**Figure 1A**).

**Figure 1.**
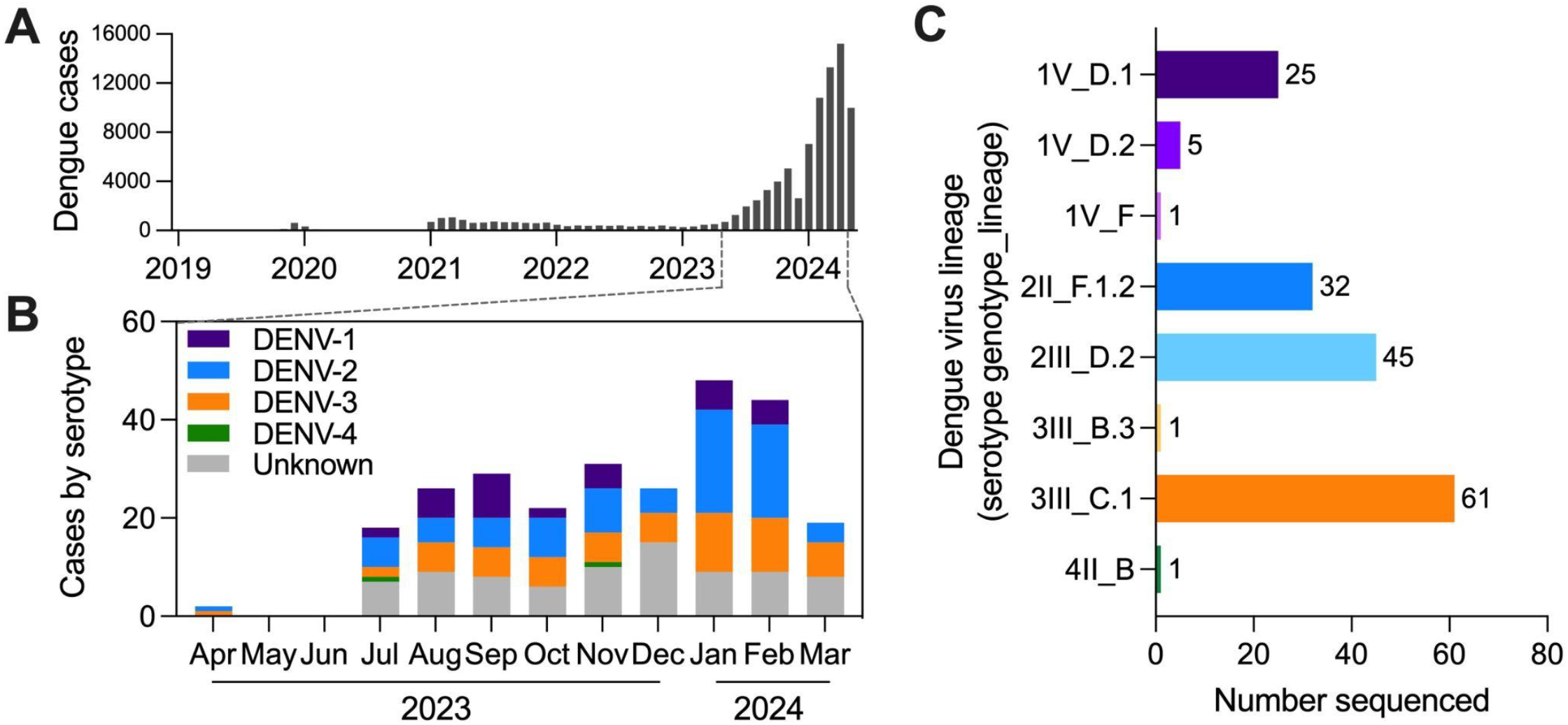
Dengue cases by virus serotype and lineage in Valle del Cauca, Colombia. (**A**) Monthly dengue cases reported by Valle del Cauca State Health Department in Colombia. Samples from confirmed dengue cases (*n*=266) sorted by DENV (**A**) serotype per month by RT-qPCR (unknown = below limits of PCR detection) and (**B**) lineage by amplicon-based sequencing (listed as serotype, genotype, and lineage).

The cause of the substantial increase in reported dengue cases is likely multifaceted. Warming temperatures caused by climate change are known to increase the transmission potential and expand the geographical range of the primary mosquito vector, *Aedes aegypti* (2). Moreover, Indian Ocean sea surface temperature anomalies, especially El Niño events, are associated with dengue epidemics (3). There was a powerful El Niño–Southern Oscillation event in 2023-24, the first since 2015-16. Moreover, new DENV introductions, perhaps due to the resumption of travel post the COVID-19 pandemic (4), could be reaching large susceptible populations. For example, DENV-3 was rarely detected in the Americas for the last 10 years before a recent introduction into the Caribbean from Asia (5,6). To investigate if there was a specific DENV serotype or lineage contributing to the recent surge in cases, we sequenced DENV infections diagnosed in Valle del Cauca, Colombia from April 2023 to May 2024.

## Methods and Results

The Valle del Cauca State Health Department in Colombia reported 966 dengue cases in 2019, 655 in 2020, 8,940 in 2021, 4,630 in 2022, 22,988 in 2023, and 56,355 cases through May, 2024 (**Figure 1A**). To determine what DENV serotypes and lineages caused some of the recent cases, we collected serum (150-500 µL) from 266 confirmed dengue cases diagnosed at Hospital Universitario del Valle (HUV) from April 2023 to May 2024, aged <1 to 77 years (average = 16 years). The samples were shipped to Yale University in New Haven, Connecticut for molecular processing. Our study was approved by the Research Ethics Committee at HUV and Yale University Human Research Protection Program (Protocol ID: 2000033281) and all participants signed an informed consent.

We extracted RNA from 140 µL of serum per sample using the QIAamp Viral RNA Mini Kit. We initially determined the DENV serotype using a multiplexed RT-qPCR assay (7) before attempting pan-serotype whole-genome amplicon sequencing with DengueSeq (8). Bioinformatic analysis, including primer trimming and consensus sequence generation, was conducted using an iVar pipeline as described in Vogels et al. (8). We assigned DENV lineages to samples with at least 5% genome completeness using the new nomenclature system (9) implemented in Genome Detective (dengue-lineages.org). We assigned serotypes to 185 of the 266 samples (70%; **Figure 1B**) and assigned lineages to 171 samples (64%; **Figure 1C**; **Appendix Table**).

We detected all four DENV serotypes from the 2023-24 samples (DENV-1 = 35, DENV-2 = 85, DENV-3 = 63, DENV-4 = 2, unknown = 81; **Figure 1B**). DENV-1, -2, and -3 were of relatively equal proportions for part of 2023, but then DENV-1 decreased as DENV-2 increased in frequency from late 2023 to early 2024. We also detected multiple lineages per serotype (except for DENV-4), with DENV-3 genotype III lineage C.1 (3III_C.1, according to (9)), DENV-2III_D.2, DENV-2II_F.1.2, and DENV-1V_D.1 being the most common (**Figure 1C**).

To further investigate the DENV lineages, we performed phylogenetic analysis using 79 sequenced samples for which we achieved >70% genome coverage (DENV-1 = 10 sequences, DENV-2 = 38, DENV-3 = 31, DENV-4 = 0). We combined our data with a background dataset downloaded from GenBank, and then downsampled the data per serotype so that we kept all sequences from Colombia, 5 per year from the other countries in the Americas, and 1 per year from each country outside of the Americas (DENV-1 = 1007 sequences, DENV-2 = 965, DENV-3 = 542). We analyzed the sequences using the Nextstrain bioinformatic and phylogenetic framework (10), including constructing maximum likelihood trees using IQTree (11) with a GTR substitution model. Our sequencing data can be found at NCBI BioProject PRJNA1132139 and the GenBank accession numbers are listed in **Appendix Table**. Our alignments, trees, and Nextstrain outputs are available at: github.com/grubaughlab/DENV-genomics/tree/master/paper_2024-CO.

Our DENV-1 phylogenetic analysis reveals the co-circulation of two distinct lineages, DENV-1V_D.1 and D.2 (**Figure 2**). Both have been previously detected in Colombia and elsewhere in South America (5,12), representing ongoing local and regional persistence and diversification of these lineages for the past ∼15-20 years.

**Figure 2.**
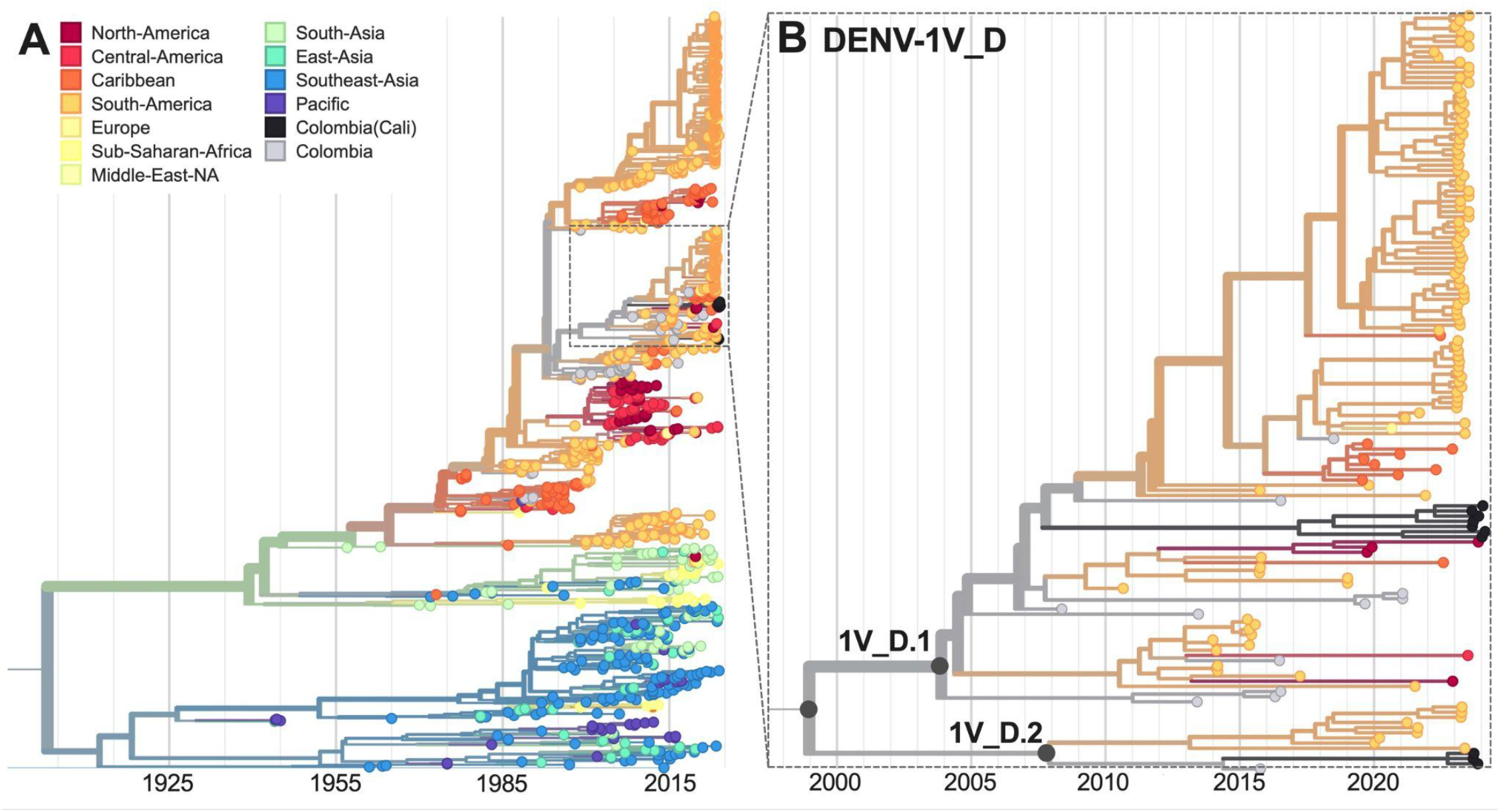
Time-resolved maximum-likelihood phylogeny of DENV-1. (**A**) Full reconstruction of 1007 DENV-1 sequences colored by sampling location. (**B**) Zoom on the DENV-1V_D clade highlighting sequences from Cali, Colombia (black).

Our phylogenetic analysis of DENV-2 presents a more complicated picture of three genetic clusters and three individual sequences dispersed among two defined lineages, DENV-2III_D.2 and DENV-2II_F.1.2 (**Figure 3**). Lineage 2III_D.2 is a descendent of the original DENV-2 genotype III (“Asian-American”) introduced in the Americas during the late 1970s that subsequently became established throughout the region, including Colombia (13). DENV-2 genotype II (“Cosmopolitan”) was recently introduced into the Americas from Asia, first detected during a dengue outbreak in Peru in 2019 (14). Detection of DENV-2II_F.1.2 in Valle del Cauca demonstrates that the emerging “Cosmopolitan” genotype can become established alongside the existing “Asian-American” genotype.

**Figure 3.**
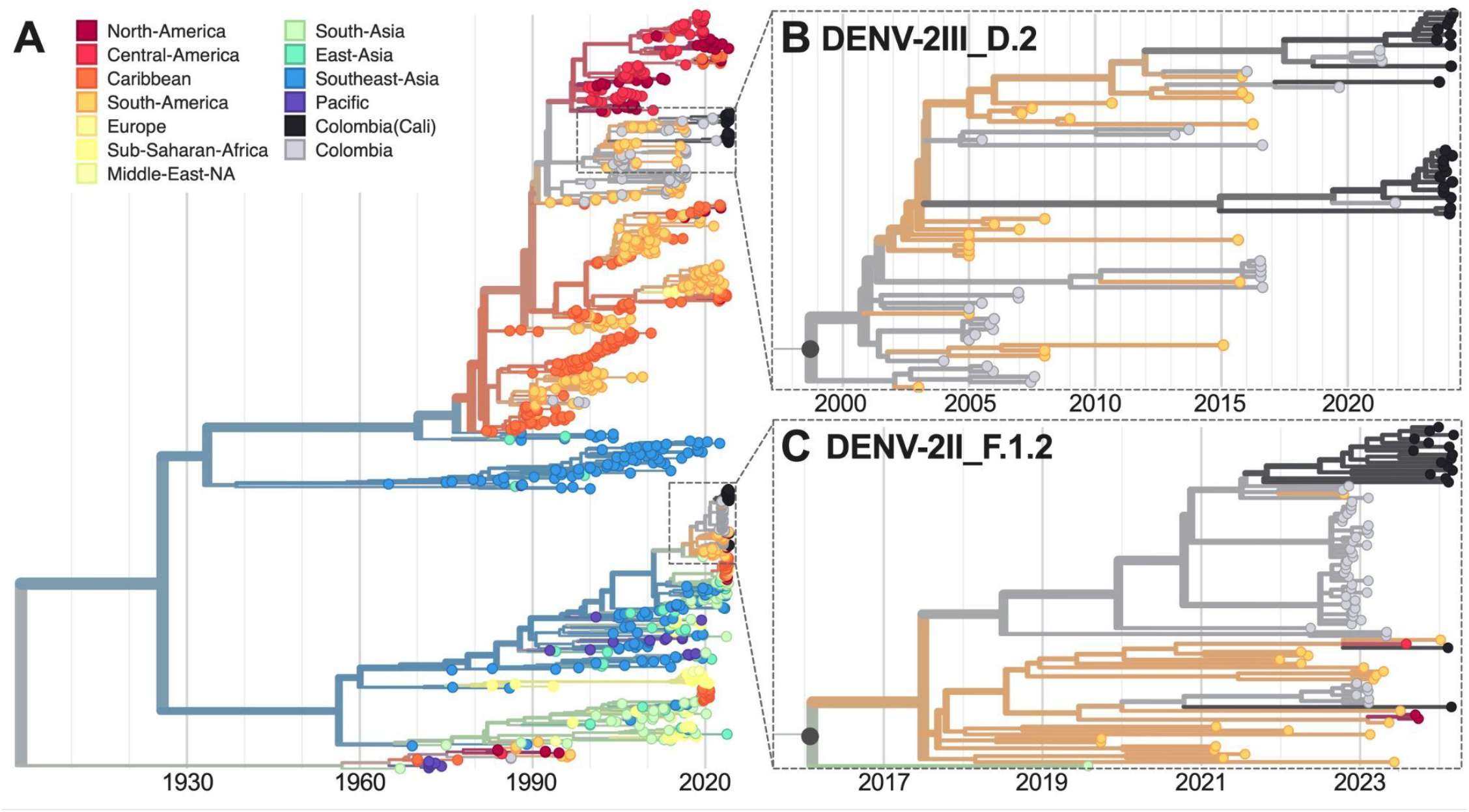
Time-resolved maximum-likelihood phylogeny of DENV-2. (**A**) Full reconstruction of 965 DENV-2 sequences colored by sampling location. Zoom on the (**B**) DENV-2III_D.2 and (**C**) DENV-2II_F.1.2 clades highlighting sequences from Cali, Colombia (black).

One hypothesis for the sudden increase in dengue cases was the introduction and rapid spread of a new DENV-3 lineage from Asia (5). DENV-3 in the Americas can often go decade or more between detections (4,6,15). Therefore, detecting an emerging lineage (DENV-3III_B.3) in the Caribbean (5), Brazil (6), Nicaragua (4), and elsewhere in the Americas was of concern. While we detected one dengue case from Valle del Cauca in January 2024 with a likely 3III_B.3 infection (18% genome coverage), 61 of 63 (97%) of DENV-3 infections were lineage 3III_C.1 (**Figure 1C**). DENV-3III_C was likely first introduced into the Americas in the early 1990s (13) and we show that it has persisted through long periods of low detection (**Figure 4**). This includes sporadic detections of 3III_C.1 in Colombia since the early 2000s. Therefore, our results suggest that populations in the Americas may be susceptible to DENV-3 in general and not just the emerging 3III_B.3 lineage.

**Figure 4.**
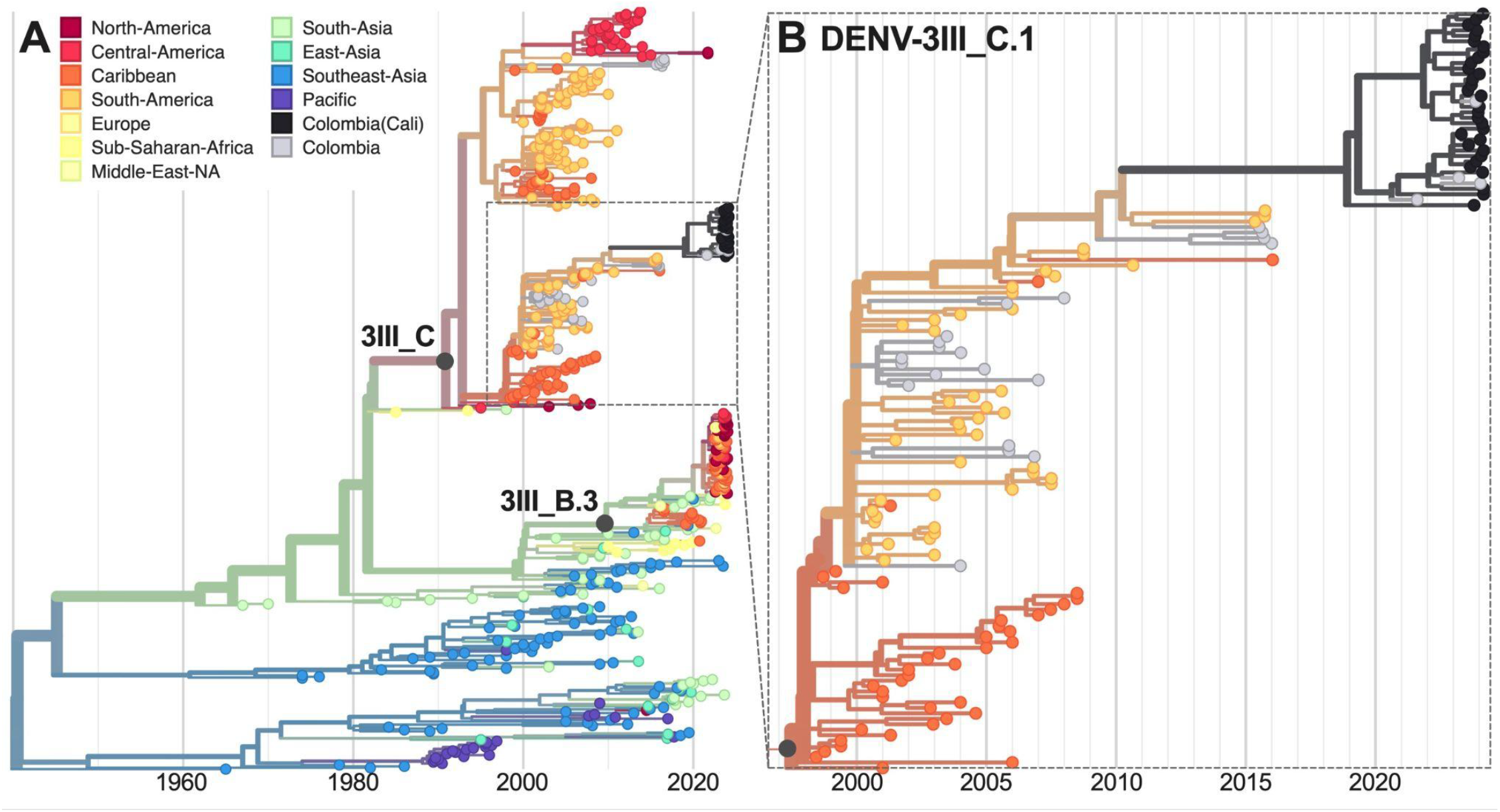
Time-resolved maximum-likelihood phylogeny of DENV-3. (**A**) Full reconstruction of 542 DENV-3 sequences colored by sampling location. (**B**) Zoom on the DENV-3III_C.1 clade highlighting sequences from Cali, Colombia (black).

## Conclusions

We investigated the DENV infections from Valle del Cauca, Colombia to determine if a specific virus serotype or lineage was in part driving the record number of dengue cases (1). We detected all four serotypes, with DENV-1, -2, and -3 sharing dominance, and at least eight separate defined lineages (9). These included multiple lineages of DENV-1 genotype V and DENV-2 genotype III (“Asian-America”) which have circulated in the Americas for ∼40 years (13), as well as the emerging DENV-2II_F.1.2 (“Cosmopolitan”) lineage. Moreover, despite the rapid spread of a new DENV-3III_B.3 lineage in the Americas (4–6), we found that the dominant DENV-3 lineage was 3III_C.1 that has been sporadically detected in Colombia for ∼20 years. Our results show that the conditions for transmission in Valle del Cauca are highly suitable for a diversity of DENV serotypes and lineages, and thus that the specific viruses are not the primary driver of the large outbreak.

## Data Availability

Our sequencing data can be found at NCBI BioProject PRJNA1132139 and the GenBank accession numbers are listed in Appendix Table. Our alignments, trees, and Nextstrain outputs are available at: github.com/grubaughlab/DENV-genomics/tree/master/paper_2024-CO.

## Acknowledgement

This publication was made possible by the National Institute of Allergy and Infectious Diseases of the National Institutes of Health (NIH) under Award Number DP2AI176740 (NDG) and an NIH Shared Equipment grant 1S10OD028669-01 award to the Yale Center for Genome Analysis. The findings and conclusions in this report are those of the author(s) and do not necessarily represent the official position of the NIH.

